# Predicting Infectivity: Comparing Four PCR-based Assays to Detect Culturable SARS-CoV-2 in Clinical Samples

**DOI:** 10.1101/2021.07.14.21260544

**Authors:** Emily A. Bruce, Margaret G. Mills, Reigran Sampoleo, Garrett A. Perchetti, Meei-Li Huang, Hannah W. Despres, David J. Shirley, Keith R. Jerome, Alexander L. Greninger, Jason W. Botten

**Affiliations:** Division of Immunobiology, Department of Medicine, Robert Larner, M.D. College of Medicine, University of Vermont, Burlington VT, 05405, USA; Virology Division, Department of Laboratory Medicine and Pathology, University of Washington, Seattle WA 98195, USA; Department of Microbiology and Molecular Genetics, Robert Larner, M.D. College of Medicine, University of Vermont, Burlington VT, 05405, USA; Faraday, Inc. Data Science Department. Burlington VT, 05405, USA; Vaccine and Infectious Disease Division, Fred Hutchinson Cancer Research Center, Seattle WA 98109, USA

**Author notes:** Correspondence (EAB), (ALG), (JWB). Department of Microbiology and Molecular Genetics, Robert Larner, M.D. College of Medicine, University of Vermont, Burlington VT, 05405, USA. EAB and MGM contributed equally to this work.

## Abstract

With the COVID-19 pandemic caused by SARS-CoV-2 now in its second year, there remains an urgent need for diagnostic testing that can identify infected individuals, particularly those who harbor infectious virus. Various RT-PCR strategies have been proposed to identify specific viral RNA species that may predict the presence of infectious virus, including detection of transcriptional intermediates (e.g. subgenomic RNA [sgRNA]) and replicative intermediates (e.g. negative-strand RNA species). Using a novel primer/probe set for detection of subgenomic (sg)E transcripts, we successfully identified 100% of specimens containing culturable SARS-CoV-2 from a set of 126 clinical samples (total sgE C_T_ values ranging from 12.3-37.5). This assay showed superior performance compared to a previously published sgRNA assay and to a negative-strand RNA assay, both of which failed to detect target RNA in a subset of samples from which we isolated live virus. In addition, total levels of viral RNA (genome, negative-strand, and sgE) detected with the WHO/Charité primer-probe set correlated closely with levels of infectious virus. Specifically, infectious virus was not detected in samples with a C_T_ above 31.0. Clinical samples with higher levels of viral RNA also displayed cytopathic effect (CPE) more quickly than those with lower levels of viral RNA. Finally, we found that the infectivity of SARS-CoV-2 samples is significantly dependent on the cell type used for viral isolation, as Vero E6 cells expressing TMRPSS2 extended the analytical sensitivity of isolation by more than 3 C_T_ compared to parental Vero E6 cells and resulted in faster isolation. Our work shows that using a total viral RNA Ct cut-off of >31 or specifically testing for sgRNA can serve as an effective rule-out test for viral infectivity.

## INTRODUCTION

Over the past year and a half, SARS-CoV-2, the etiologic agent of COVID-19, has caused extraordinary disruption on a global scale. While sensitive and accurate tests were developed early in the pandemic to detect the presence of SARS-CoV-2 RNA, it has become clear that many patients continue to test positive for weeks after the resolution of symptoms [1, 2]. In addition, recent work has shown that the period of RNA positivity can substantially outlast the period of time in which infectious virus is present in a patient [3]. With current levels of global spread, and the quarantine and personal protective equipment (PPE) requirements required following positive tests, there is an urgent need in this and potential future pandemics to determine which individuals testing positive by RT-PCR are still capable of transmitting virus to others. The gold standard to determine infectivity involves culturing patient samples on a susceptible cell line and confirming the presence or absence of infectious SARS-CoV-2. This approach is not practical for individual clinical diagnoses, however, as it requires a biosafety level (BSL)-3 laboratory, is unsuitable for high throughput processing, and has no FDA-authorized viral isolation diagnostic test available. Culturing of patient samples has indicated that most patients are infectious only until about 10 days after symptom onset [4, 5], but in rare cases infectivity can persist much longer [6]. Determining when RT-PCR-positive patients are no longer infectious and can therefore be released from quarantine is a question of great clinical relevance and personal importance for many patients and medical professionals.

SARS-CoV-2, like other coronaviruses, produces a nested set of subgenomic RNA (sgRNA) species during viral replication that are required to express the viral structural proteins. Each sgRNA is composed of the 5’ leader sequence from the whole genome appended to the reading frame for one gene by discontinuous transcription, with the short Transcription Recognition Sequence (TRS) separating them [7, 8]. This process brings sequences that are tens of kilobases apart in the genome to be only tens of bases apart in the sgRNA, allowing them to be identified by routine RT-PCR. Because sgRNAs are generated only during replication, the detection of sgRNAs in patient samples by RT-PCR has been used as a marker of active viral replication [4, 9-11], and the absence of sgRNA has been used in notable circumstances to clear patients from quarantine requirements [12]. There are conflicting reports regarding the clinical utility of using the presence of sgRNA as a predictor of infectivity. Some studies promote its use [10, 11] while others (including those using the Wölfel-sgE primer-probe set) deemed it unsuitable for predicting the presence of infectious virus [5, 13, 14]. However, the presence of sgRNA has been used to successfully distinguish input challenge virus from actively replicating virus, particularly in non-human primate models of SARS-CoV-2 vaccination and challenge [15-19]. Similarly, there has been recent interest in using the presence of negative-strand RNA, a direct product of viral RNA replication, to identify patients with active viral replication [20]. Herein, we sought to develop a sgRNA assay that would overcome possible limitations of existing sgRNA primer-probe sets and test whether sgRNA detection can effectively identify clinical samples harboring infectious SARS-CoV-2.

## RESULTS

### The Mills assay to measure SARS-CoV-2 subgenomic (sg)RNA is specific and sensitive

Existing primer sets to detect sgRNAs result in longer amplicons than used for comparison genomic RNAs [4], which we hypothesized might reduce sensitivity, and used probes entirely within gene coding regions, which might reduce specificity of the probes for sgRNAs. Accordingly we designed an alternative primer-probe set (termed Mills-sgE) that targets the sgE mRNA using a forward primer in the leader sequence of the genome, a reverse primer near the 5’ end of the E gene, and a probe that binds to the TRS junction between the leader and E gene in the sgRNA (Fig. 1).

**Figure 1.**
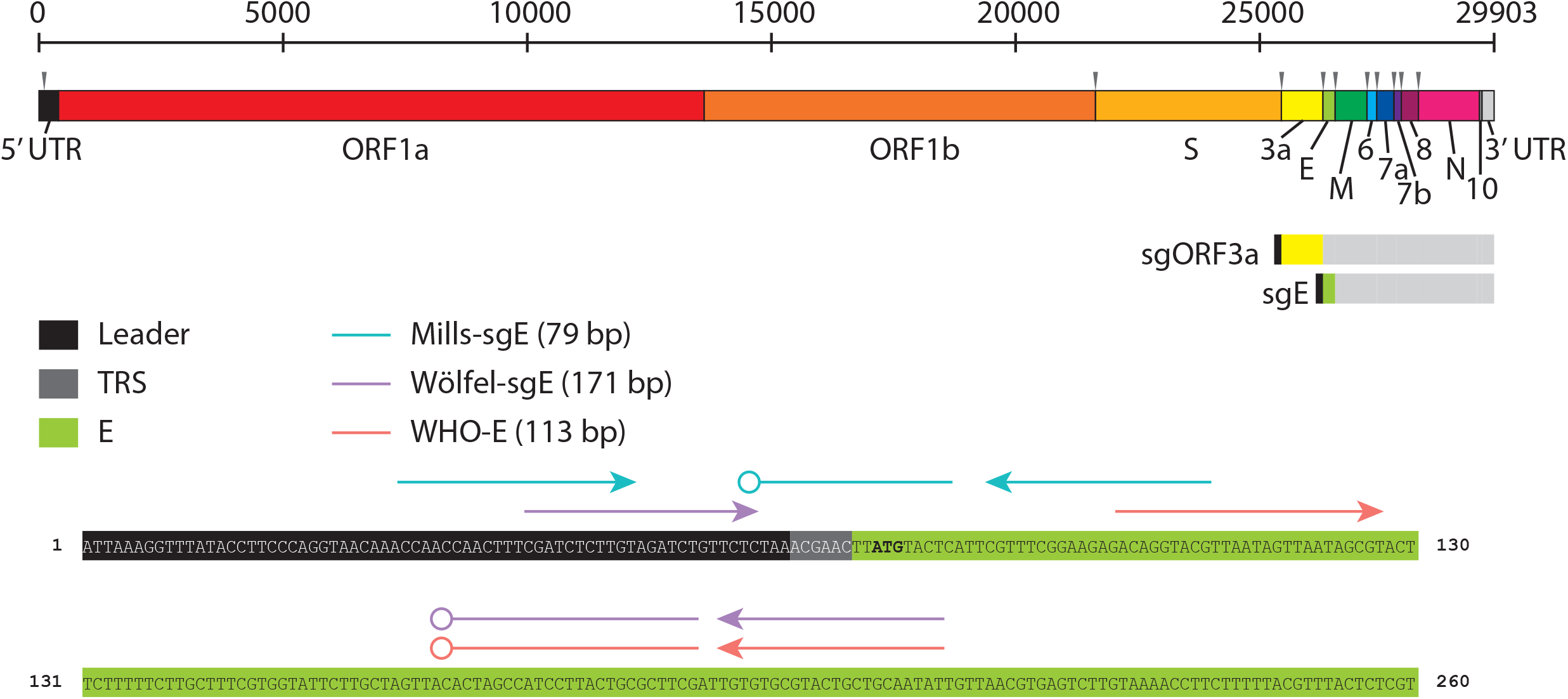
SARS-CoV-2 genome structure and Mills-sgE subgenomic primer/probe set design. The organization of the SARS-CoV-2 genome is illustrated, including each coding region and untranslated regions (UTR) (names indicated below) as well as Transcription Recognition Sequences (TRS, grey arrowheads above). Subgenomic RNAs for each viral structural protein (ORF3a and E shown) consist of the Leader (L) sequence from the 5’ UTR appended to the 5’ end of the coding region separated by the TRS. The location of primer-probe sets used in this study to detect E (WHO E gene [27]) and sgE (Wölfel-sgE [4] and Mills-sgE) are identified by lines with arrowheads (primers) and open circles (probes), respectively, above the partial sequence of sgE.

We confirmed the accuracy of the Mills-sgE assay by measuring Mills-sgE in our negative extraction control (HeLa cells), as well as clinical samples that had tested negative by Hologic Panther Fusion; no amplified product was detected. We further confirmed the specificity of the assay by testing it on excess clincial specimens from the University of Washington clinical virology laboratory (UWVL) that had previously been measured to have high copy number of 12 different respiratory viruses, including adenovirus (AdV), bocavirus (BoV), influenza A (IAV), influenza B (IBV), metapneumovirus (MPV), parainfluenzavirus1-4 (PIV1, PIV2, PIV3, PIV4), rhinovirus (RhV), respiratory syncytial virus (RSV) and 24 samples positive for other human coronaviruses. No nonspecific amplification of other human viruses was detected with the Mills-sgE primer set. In addition, the Mills-sgE assay did not amplify the commercially-available AccuPlex synthetic SARS-CoV-2 genome, indicating that the assay specifically identified subgenomic RNA and not full-length genomic RNA from SARS-CoV-2 (Supplemental Table S1).

The sensitivity of the Mills-sgE assay was measured with an *in vitro* transcribed fragment of sgE that had been quantified by RT-digital drop (dd)PCR. Dilutions of the stock were measured in quadruplicate to determine an initial limit of detection (LoD), and were then confirmed with 20 replicates at each concentration, where the LoD was defined as the last dilution to detect at least 19/20 positives. This identified the LoD at 1.1 copies/μL or approximately 5 sgE copies/reaction (Table 1).

**Table 1.** Mills-sgE Limit of Detection.

| At LoD |  |  | Beyond LoD |  |  | LoD<br>(copies/rxn) |
| --- | --- | --- | --- | --- | --- | --- |
| Concentration<br>(copies/μL) | Mean<br>C<sub>T</sub> | Positives<br>Detected | Concentration<br>(copies/μL) | Mean<br>C<sub>T</sub> | Positives<br>Detected |  |
| 1.06 | 37.6 | 19/20 (95%) | 0.53 | 38.3 | 15/20 (75%) | 5.3 |

### Kinetics of SARS-CoV-2 infection in two Vero E6 cell lines

In order to identify clinical samples that contained infectious virus, we first tested two cell lines for their ability to support SARS-CoV-2 infection. Vero E6 cells are a standard cell line used for viral isolations (for SARS-CoV-2 and many other viruses) as they are typically permissive to viral infection due to their inability to produce IFN alpha or beta. However, they do not express the TMPRSS-2 protease present in lung cells which are the natural targets of SARS-CoV-2 and thus do not fully recapitulate the physiological entry pathway [21, 22]. In contrast, Vero E6 cells that stably express TMPRSS-2 facilitate entry at the cell surface and are a tractable cell culture model. We therefore investigated the kinetics of viral RNA and infectious virus production in Vero E6 and Vero E6-TMPRSS2 cells infected with SARS-CoV-2 strain WA1. Cells were infected at an MOI of 0.001 and supernatants and cell lysates were collected throughout the time course of infection. In Vero E6-TMPRSS2 cells, viral growth reached peak titer 1 day post infection (dpi) and declined to near undetectable levels by 4 dpi, with nearly complete CPE by 3 dpi (Fig 2A). Viral replication occurred with slightly delayed kinetics in Vero E6 cells, and though the peak levels of virus produced were only slightly lower than in Vero E6-TMPRSS2 cells, more than four logs of virus remained at eight days post infection (Fig 2B). This delay in virus and RNA production likely reflects delayed viral entry leading to slower viral spread, and is in keeping with the rate of CPE observed in the two cell lines over the course of infection; while CPE was visible by 3 dpi in the Vero E6 cells, it was not complete even at 8 dpi.

**Figure 2.**
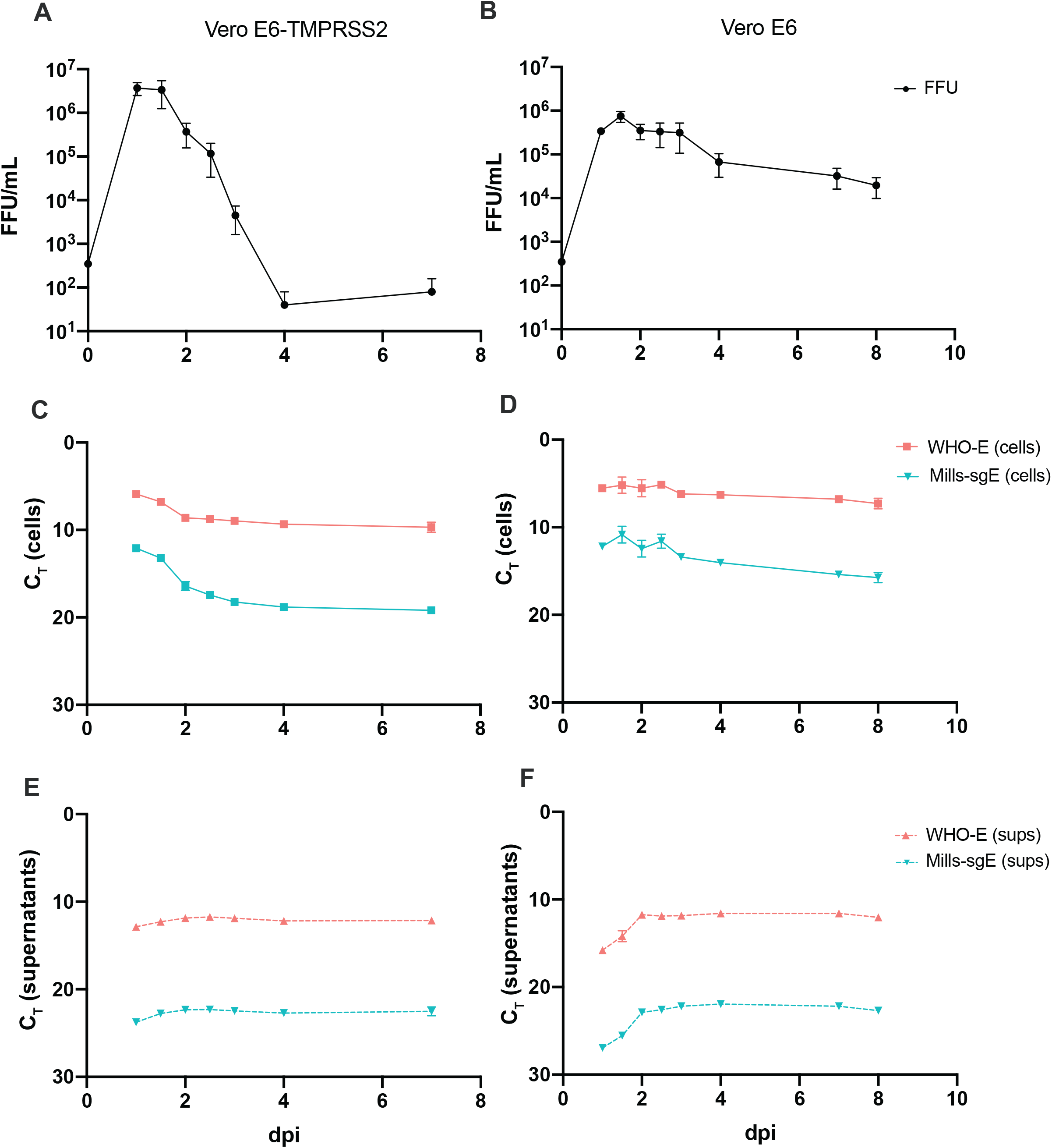
Kinetics of SARS-CoV-2 growth in Vero-E6 and Vero E6-TMPRSS2 cells. (A, C, E) Vero E6-TMPRSS2 or (B, D, F) Vero-E6 cells were infected with SARS-CoV-2 WA1 at an MOI of 0.001. Supernatants were collected twice daily for two days, daily for an additional two days, and cells and supernatants were collected for a final timepoint at day 7 (for Vero E6-TMPRSS2 cells) or day 8 (for Vero E6 cells). A, B) Viral titer was determined by immune-focus forming assay (focus-forming units [FFU]/mL), day 0 values are calculated based on known titer of inoculum. RNA levels were determined by RT-PCR from cells (C, D) or viral supernatants (E, F) for each timepoint using the indicated primer-probe sets. Note that C_T_ is plotted inversely on the Y axes, so that lower RNA concentrations are shown lower on the axes. All data points represent four technical replicates from two independent experiments and are plotted as mean with error bars plotted as +/-SEM. Limit of detection for infectious titer is plotted as the minimum y value.

We investigated the kinetics of viral RNA expression in both cultures. As expected, viral RNA detected by a primer set for the Envelope (E) gene used by the World Health Organization (referred to as WHO-E here), which detects primarily positive-strand genomic but also subgenomic and negative-strand RNA, was observed in both cells and supernatants. Vero E6 cells supported higher levels of intracellular viral RNA production, along with their sustained production of infectious virus (Fig 2D), than did Vero E6-TMPRSS2 cells (Fig 2C). In both cell lines, viral RNA levels within cells declined from their peak during the first two days of infection before plateauing at a stable level for the subsequent five days (Fig. 2C, D). In supernatants, there was a slight increase in viral RNA levels observed during the first two days of infection, then levels remained constant throughout the next five days (Fig 2E, F). As expected, subgenomic E RNA expression detected by the Mills-sgE primer-probe set was a fraction (6-9 C_T_ less, roughly 1/50 to 1/500) of total viral RNA that was detected by WHO-E in cells (Fig. 2C, D). Notably, subgenomic E RNA was detected in cell-free supernatants (10-11 C_T_ less than that detected with WHO-E, roughly 1/1000 to 1/2000) (2E, F), and the ratio of subgenomic RNA to total viral RNA remained surprisingly similar throughout the time course.

In summary, SARS-CoV-2-induced CPE and infectious virus production was accelerated and of higher magnitude in Vero E6-TMPRSS2 cells versus standard Vero E6 cells. And regardless of cell line, the viral RNA species measured in cells or virions remained relatively stable over the time course, even as infectious virus titers waned. Notably, sgRNA was consistently found in cell-free supernatants, suggesting it may be packaged into virions.

### Stability of infectious SARS-CoV-2 particles and viral RNA species

We observed that SARS-CoV-2 RNA species persist for much longer than infectious virus in cell culture time course experiments (Fig. 2). In particular, we noted an ∼5 log drop in live virus over a three-day period in Vero E6-TMPRSS2 cells while RNA levels remained stable in supernatants for at least seven days (Fig. 2A, C, E), and likely considerably longer given no decrease was observed in that time. We hypothesized that this disparity could be due to differences in the stability of live virus versus viral RNAs. To test this, we subjected samples to various real-world handling conditions and measured the stability of both infectious SARS-CoV-2 virus and RNAs within those samples. Aliquots of pooled patient specimens at three different concentrations were subjected to 4°C storage for periods between 1 and 14 days, or for -80°C storage interrupted by up to 5 freeze-thaw cycles (Fig. 3A, B). Similarly, viral stocks of SARS-CoV-2 WA1 were subjected to storage at 4°C, room temperature, or 37°C, or, for -80°C storage, interrupted by up to 6 freeze-thaw cycles (Fig. 3C, D). Both infectious virions and sgE RNA showed impressive stability through storage at 4°C (Fig. 3B, D), storage at room temperature (Fig. 3D), and through repeated freeze-thaw cycles (Fig. 3A, C), though viral infectivity declined precipitously at 37°C (Fig. 3D). Only the lowest concentration of pooled patient sample showed any decrease in sgE concentration through refrigerated storage (Fig. 3B), or through repeated freeze-thaw cycles (Fig. 3A). Thus, it appears that the disparity observed between levels of infectious virus and viral RNA seen at later points in our viral growth curves may reflect a drastic difference in stability at 37°C, with viral RNA far more robust than live virus at this temperature. Collectively, these results suggest that viral RNA and infectious virus contained in COVID-19 clinical samples likely remain stable under a variety of real-world field conditions, including freeze thaws or extended storage at 4°C or room temperature.

**Figure 3.**
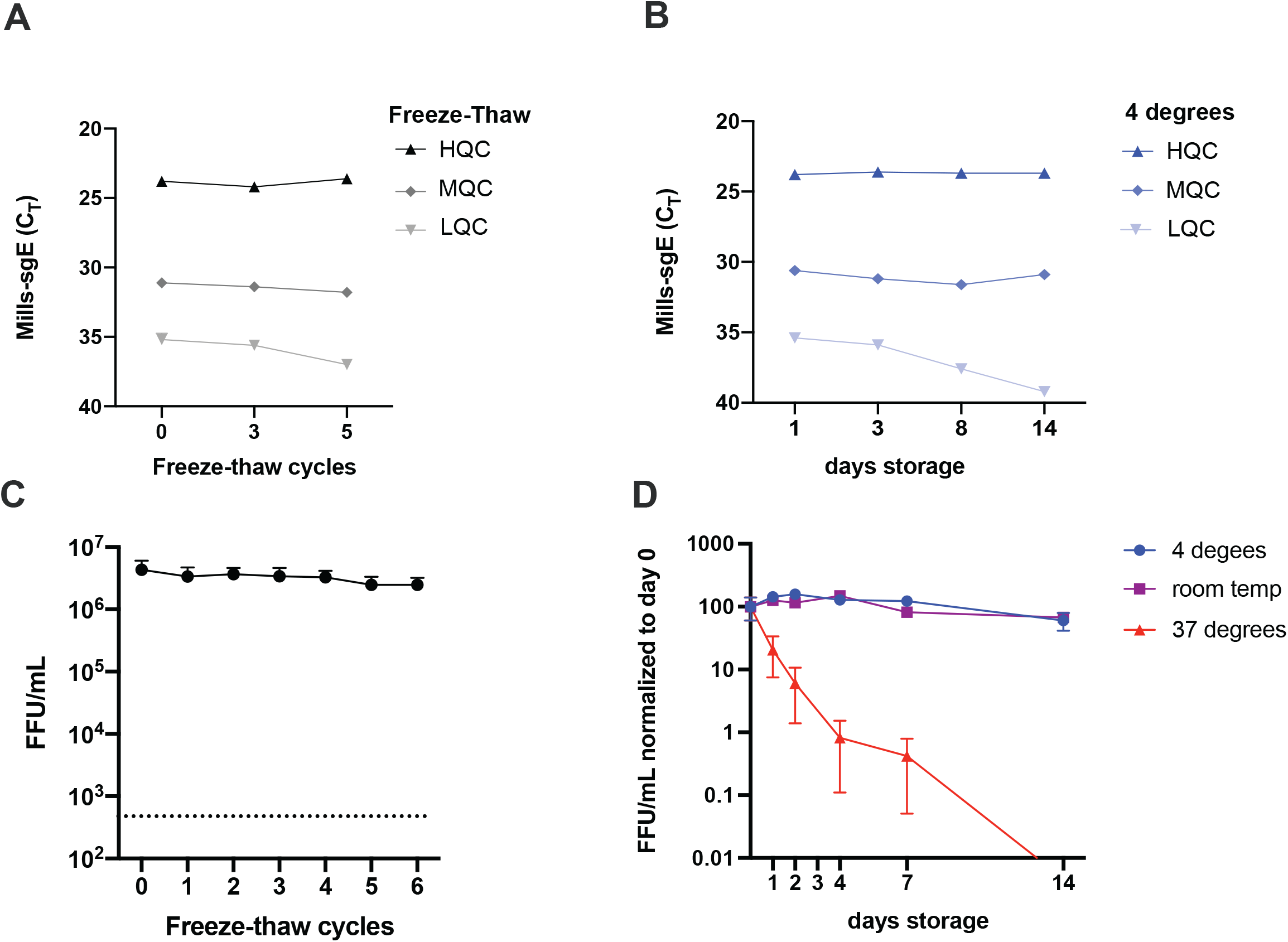
Stability of sgRNA and viral infectivity during freeze-thaw cycles and storage. (A, B) Pooled patient samples at high (5.7×10^4^ copies), medium (3.6×10^2^ copies) or low (2×10^1^ copies) concentrations, or (C, D) stocks of SARS-CoV-2 WA1 were subjected to -80°C storage interrupted by (A, C) successive freeze-thaw cycles or (B, D) storage at 4°C, room temperature or 37°C for the indicated time periods. N=1 for A, B. For C and D datapoints are from 3 different viral stocks titered in duplicate and plotted as mean with error bars plotted as +/-SEM. For C raw values are plotted while values are normalized to starting titer for D. Limit of detection for infectious titer in C is indicated by a dashed line.

### Correlation between detection of subgenomic RNA and isolatable virus in clinical samples

To determine whether detection of sgE by the Mills-sgE primer/probe set could accurately predict infectivity, we selected 126 clinical specimens across a range of clinical RT-PCR cycle threshold (C_T_) values for WHO-E (range 12.3-37.5, IQR 22.7-32.8) for viral isolation and subsequent analysis. We observed that the Vero-E6 TMPRSS2 cells were more permissive than the parental line (1.9 times the odds of being culturable compared to Vero E6 cells, p=0.02, 95% CI for OR [1.09, 3.36] by Fisher Exact test), with infectious virus detectable in 54 versus 32 clinical samples, respectively (Table 2). This difference in isolation was generally due to enhanced recovery among viruses with lower levels of SARS-CoV-2 RNA (95% of samples that isolated had a WHO-E C_T_ of <30.5, vs <27.1 for Vero E6 cells). Only 5% of samples that failed to isolate in either Vero E6 or Vero E6-TMPRSS2 cells had a C_T_ of less than 22.6, in agreement with previous reports [4, 5, 23-26]. We used our Mills-sgE assay to specifically determine sgE levels in the same 126 clinical samples used for viral isolations (Fig. 4A, B). The assay was able to detect template in all clinical samples in which infectious virus was isolated by either cell line, corresponding to a 100% negative predictive value for isolatable virus. As we observed in the cell culture time-course, however, there were many clinical samples in which we detected sgE but not isolatable virus (of the 93 samples in which we detected sgE we were only able to isolate virus from 52), resulting in a positive predictive value (PPV) of 56%.

**Table 2.** Viral RNA levels and the presence of culturable virus in clinical samples.

| Sample<br>Category<sup>a</sup> | Average Viral RNA Load (C<sub>T</sub>)<sup>b</sup> |  |  |  | Viral Isolation |  |
| --- | --- | --- | --- | --- | --- | --- |
|  | WHO-E<sup>c</sup> | Wölfel-sgE<sup>d</sup> | Mills-sgE<sup>e</sup> | Negative<br>strand E<sup>f</sup> | Vero-E6<br>TMPRSS2 | Vero-E6 |
| **High (C<sub>T</sub> <20)** | 16.6 | 22.6 | 24.2 | 27.0<br>(21 measured) | 21/22 (96%) | 21/22 (96%) |
| **Intermediate**<br>**(C<sub>T</sub> 20-30)** | 26.2 | 32.2<br>(2 NDET<sup>g</sup>/<br>54 measured) | 33.8<br>(1 NDET/<br>54 measured) | 33.5<br>(21 NDET/<br>26 measured) | 30/54 (56%) | 11/54 (20%) |
| **Low (C<sub>T</sub> >30)** | 33.8 | 37.0<br>(38 NDET/<br>50 measured) | 38.2<br>(32 NDET/<br>50 measured) | 37.7<br>(37 NDET/<br>38 measured) | 3/50 (6%) | 0/50 (0%) |
| **Total** | 27.5 | 30.4<br>(40 NDET/<br>126 measured) | 32.4<br>(33 NDET/<br>126 measured) | 28.6<br>(58 NDET/<br>85 measured) | 54/128<br>(42%) | 32/128<br>(25%) |

<sup>a</sup>Based on the values determined by the WHO-E primer-probe set in original clinical samples.
<sup>b</sup>Average is of sample values with a detectable C<sub>T</sub>.
<sup>c</sup>Levels of E viral RNA (not strand or RNA species specific), as determined by the WHO-E primer-probe set [27].
<sup>d</sup>Subgenomic (sg)E RNA levels, as determined by the primer-probe set described by Wölfel et al [4].
<sup>e</sup>sgE RNA levels, as determined by the Mills primer-probe set described in this study.
<sup>f</sup>Negative-strand E RNA levels as determined by the primer-probe strategy described in [42].
<sup>g</sup>NDET: not determined

**Figure 4.**
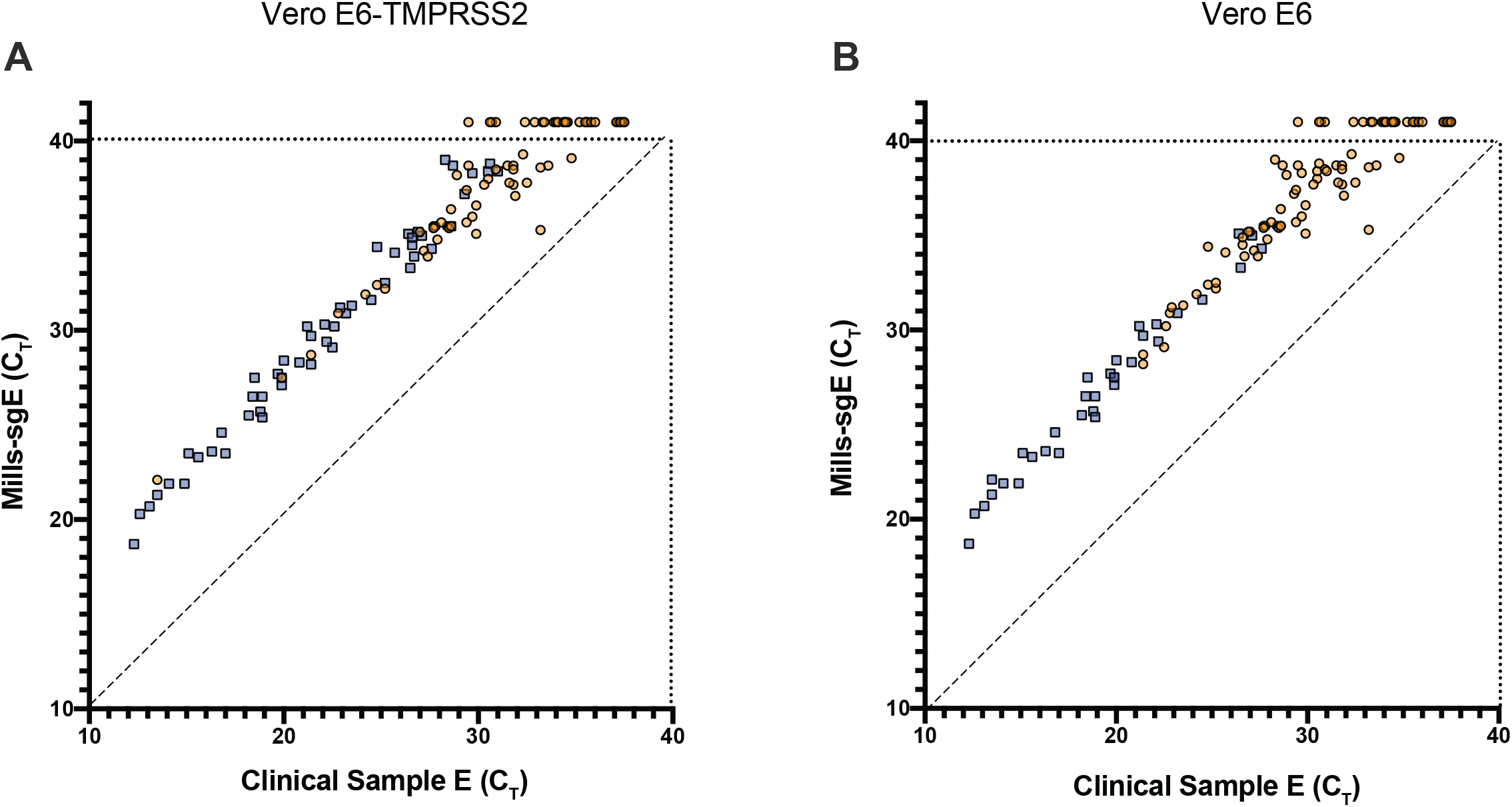
Viral isolation from SARS-CoV-2 RT-PCR-positive clinical samples in Vero-E6 and Vero-E6 TMPRSS2 cell lines. (A) Vero E6-TMPRSS2 or (B) Vero E6 cells were inoculated with a total of 126 clinical NP swab samples representing a range of viral RNA levels, both total E RNA (Clinical Sample E C_T_ detected by WHO-E primer/probe set) and subgenomic E RNA (C_T_ detected by the Mills-SgE primer set developed in this study). Culture supernatants were harvested when cells displayed CPE or at day 7 if no CPE was observed earlier, clarified, and presence (blue square) or absence (yellow circle) of SARS-CoV-2 determined by immuno-focus assay.

To investigate the relationship between levels of viral RNA and infectivity, we determined the number of days required for virus in each clinical sample to replicate to the point that cytopathic effect (CPE) was visible on Vero E6/Vero E6-TMPRSS2 monolayers. Greater initial viral RNA levels resulted in faster viral growth in both cell lines, suggesting that viral load as measured by standard RT-PCR of SARS-CoV-2 RNA is a reasonable predictor for the presence of infectious virus in patient NP samples (Fig. 5). Vero E6-TMPRSS2 cells appear more permissive than parental Vero E6 cells to SARS-CoV-2 by this measurement as well, with the majority of samples causing CPE during day 1-3 in Vero E6-TMPRSS2 cells. The same samples inoculated in parallel on Vero E6 cells took 3-7 days to cause the same level of cell death (or failed to replicate altogether). Viral titers at the endpoint of these growth curves (harvested when unambiguous CPE was observed, generally when ∼50% of cells were dead) spanned a wide range, with final titers as low as 10^3^ focus forming units (FFU)/ml and as high as 10^8^ FFU/ml in both Vero E6 and Vero E6-TMPRSS2 cells (Fig. 5). Because the samples used to measure correlates of infectivity were clinical samples, collected for research use only after clinical testing for total E RNA, the storage conditions to which they were subjected prior to testing was dictated by the requirements for SARS-CoV-2 RNA detection and capacity of the clinical laboratory rather than by what was optimal for viral isolation. The time period between sample collection and freezing for transport to the BSL-3 facility for viral isolation ranged from 1.5 to over 8 days, and part of that storage may have been at room temperature with the rest of the storage at 4°C. In our hands however, freeze-thaw cycles and room temperature storage of high titer stocks are not associated with any significant loss in infectivity (Fig. 3), suggesting that variation in clinical storage conditions was unlikely to result in a decrease in infectious virus.

**Figure 5.**
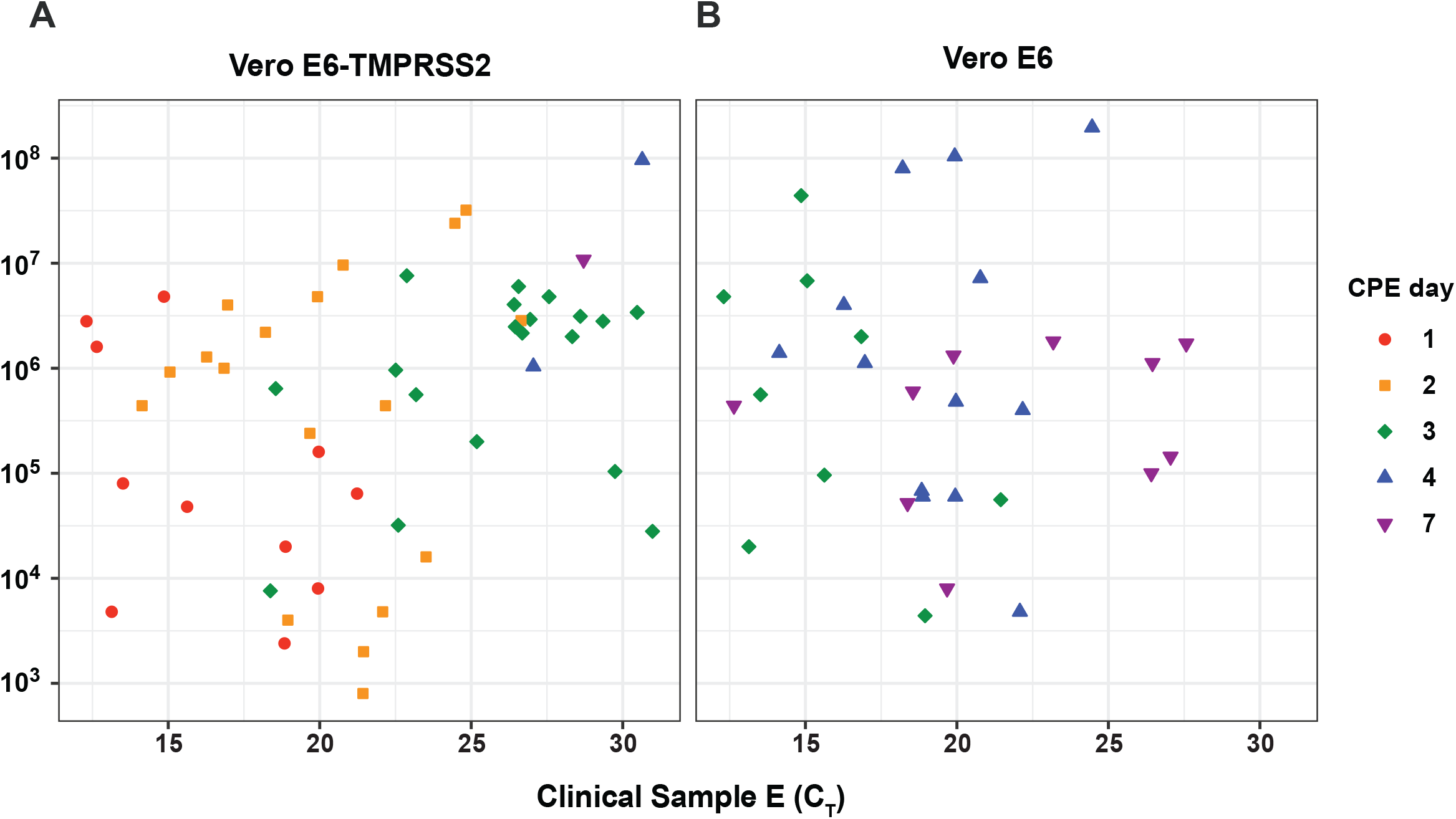
Kinetics of SARS-CoV-2 isolation. Supernatants from the panel of infected (A) Vero E6-TMPRSS2 and (B) Vero E6 cells shown in Fig. 4 were harvested when CPE was observed and the viral titer (FFU/mL measured by focus forming assay) was determined for the final timepoint of each sample that successfully isolated. Samples are colored according to the day CPE was observed and the sample was harvested. Total E RNA C_T_ of the original clinical sample used for viral isolation is plotted on the x axis. Data shown is from the subset of samples that successfully isolated in each cell line. Limit of detection for infectious titer is 400 FFU/mL.

### Alternatives to Mills-sgE

Because of the wide interest in identifying correlates of infectivity in patient samples, we compared other commonly-used methods to Mills-sgE. First, we used a primer/probe set designed by Wölfel and colleagues [4] to detect sgE in each of the clinical samples used for viral isolations. This set (termed Wölfel-sgE here) consists of a forward primer in the 5’ leader sequence of the genome, together with the reverse primer and probe from the WHO-E gene set developed by the same group [27] (Fig. 1). We observed a broad correlation between the levels of Wölfel-sgE and the level of Mills-sgE in clinical specimens, and thus with the presence of infectious virus (Fig 6A). For the 83 clinical samples for which both Wölfel and Mills CT values were successfully collected, on average the Mills C_T_ was 1.58 cycles higher than Wölfel (IQR 1.36 to 1.82). These findings are consistent with the hypothesis that the Mills-sgE primer set detects fewer SARS-CoV-2 RNA species than the Wölfel-sgE primer set due to the fact that the Wölfel probe is fully in the E coding sequence while the Mills probe spans the junction. Interestingly, for ten samples near the limit of detection (three of which contained isolatable virus), the Mills-sgE primers were able to amplify RNA that was undetectable using the Wölfel-sgE primers (Fig 4A, Fig. 6A, Supplemental Table S2). This is in line with recent studies that have reported that sgRNA levels (using Wölfel-sgE primers) are insufficient to detect the presence of infectious virus from all samples in which it can be isolated [5, 13]. While we do not have a complete explanation at this time for this observation, the Mills-sgE primer set appears to be more accurate at ruling out the presence of infectious virus than the Wölfel-sgE primer set, consistent with the hypothesis that the Mills-sgE primer set is more efficient at detecting true sgE RNAs because of shorter amplicon length.

**Figure 6.**
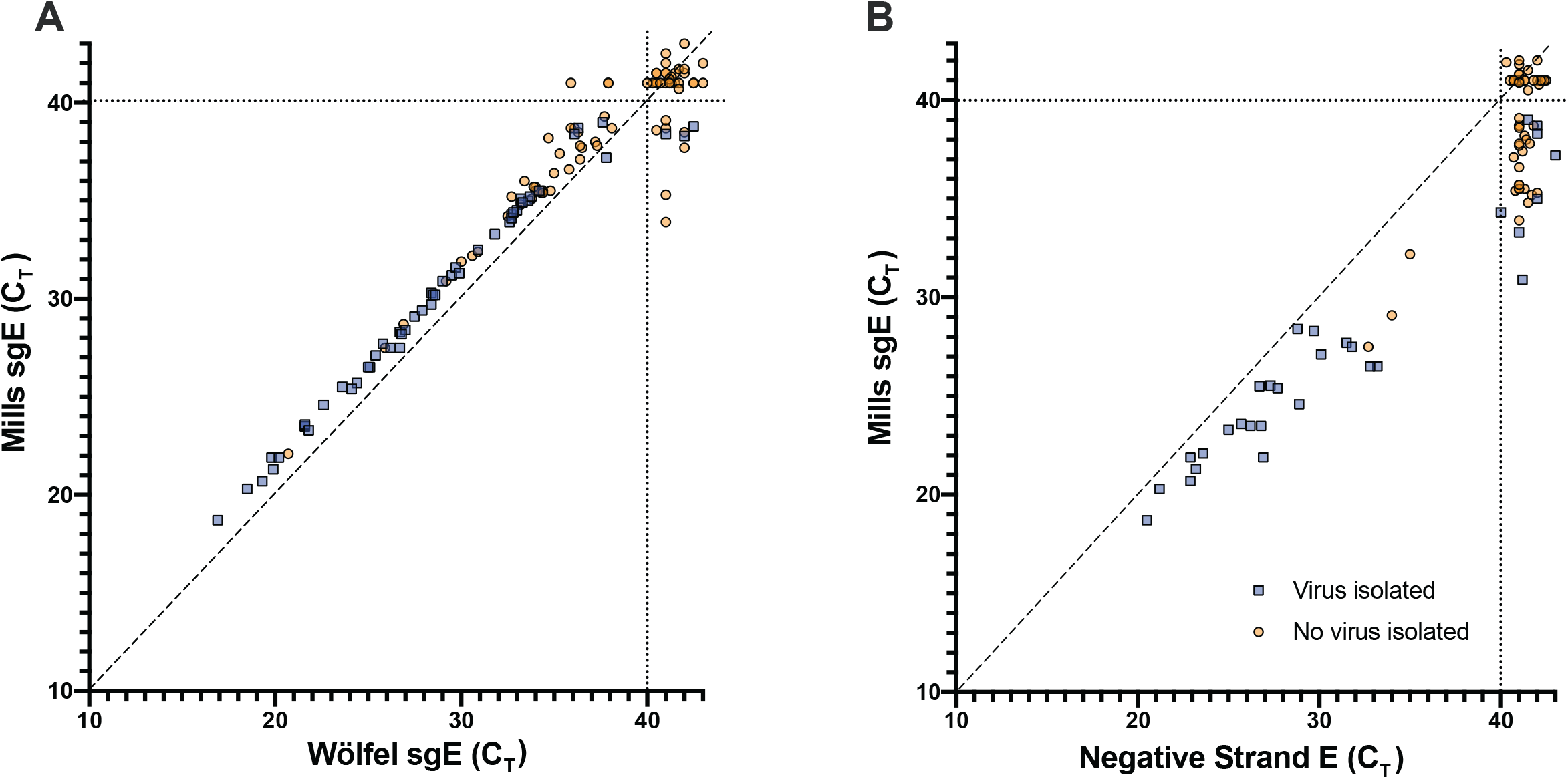
Detection of sgE by Wölfel-sgE primer set, or negative-strand RNA in clinical samples. (A) The amount of subgenomic E RNA in the panel of clinical samples used to infect Vero E6-TMPRSS2 cells in Fig. 4 was measured using a previously-published primer-probe set (Wölfel-sgE) [4] as well as a set developed for this study (Mills sgE). (B) For a subset of 85 NP swab samples for which sufficient material remained, SARS-CoV-2 negative-strand E RNA levels were determined by strand-specific RT-PCR with tagged primers. Culture supernatants were harvested when cells displayed CPE or at day 7 if no CPE was observed earlier, clarified, and presence (blue square) or absence (yellow circle) of infectious SARS-CoV-2 determined by immuno-focus assay.

Because of recent interest in using negative-strand RNA to identify patients with active viral replication [20], we tested all the patient samples for which we had sufficient volume for the presence of negative-strand RNA and compared this to our previously-obtained measures for each sample (Supplemental Table S2, Fig. 6B). We found that negative-strand E was produced at lower levels than sgE in all samples (1-7 C_T_ less than Mills sgE, roughly ½ to 1/128). For low C_T_ samples, negative-strand E has a similarly good positive predictive value (PPV) to Mills-sgE or Wölfel-sgE (Fig. 6A), with very few clinical samples that were positive for negative-strand E from which we could not isolate virus. However, the amount of negative-strand E appears to be even lower than that of sgE (8-14 C_T_ less than E, roughly 1/250 to 1/16000). Accordingly, of the higher-C_T_ samples we were able to test for negative-strand E, eight were not detected (NDET) for negative-strand E but had isolatable virus (Fig. 6B). It is clear that detection of negative-strand E RNA is substantially less sensitive than detection of sgRNAs by the Mills-sgE primer set.

To compare the performance of the four diagnostic approaches tested (WHO-E, Wölfel-sgE, Mills-sgE, negative-strand E) we generated receiver operating characteristic (ROC) curves, to display the performance of each primer/probe set (true-positive and false-positive rates) over the range of C_T_ values from which we attempted viral isolation. As the Vero E6-TMPRSS2 cells were the most sensitive cell line for viral isolation we used the viral growth status for each clinical sample in this cell line to determine the suitability of each primer-probe set to correctly identify infectious samples. Of the markers considered, C_T_ (E) was the most sensitive marker for infectivity (Fig. 7). C_T_ (Wölfel-sgE) and C_T_ (Mills-sgE) had similar infectivity sensitivity-specificity profiles, which were both slightly inferior to C_T_ (E). Because fewer samples were tested for negative-strand E (due to limitations in available surplus sample material), we generated a second set of ROC curves using just the data from these samples. C_T_ (negative-strand E) had a similar infectivity sensitivity-specificity profile to those for both C_T_ (Wölfel-sgE) and C_T_ (Mills-sgE) (Fig S1). However, with the protocols used here, both the negative-strand E and Wölfel-sgE assays were unable to detect 95% of infective samples (NPV = 87.9% and 92.5% respectively). Assays for WHO-E and Mills-sgE both had 100% NPV, detecting 100% culturable samples in Vero-TMPRSS2 cells at C_T_ values of 31.0 and 38.7, respectively. Using a C_T_ cut-off of 40, the assay for Mills-sgE offered a much lower false-positive rate than the assay for WHO-E: only 56% of the samples that were positive for E and did not culture were detectable by the Mills-sgE primer set (PPV = 55.9% vs 41.3% for E > C_T_ 40). But overall, the best test for infectivity was a C_T_ cut-off of 31.0 for WHO-E (PPV = 61.2%).

**Figure 7.**
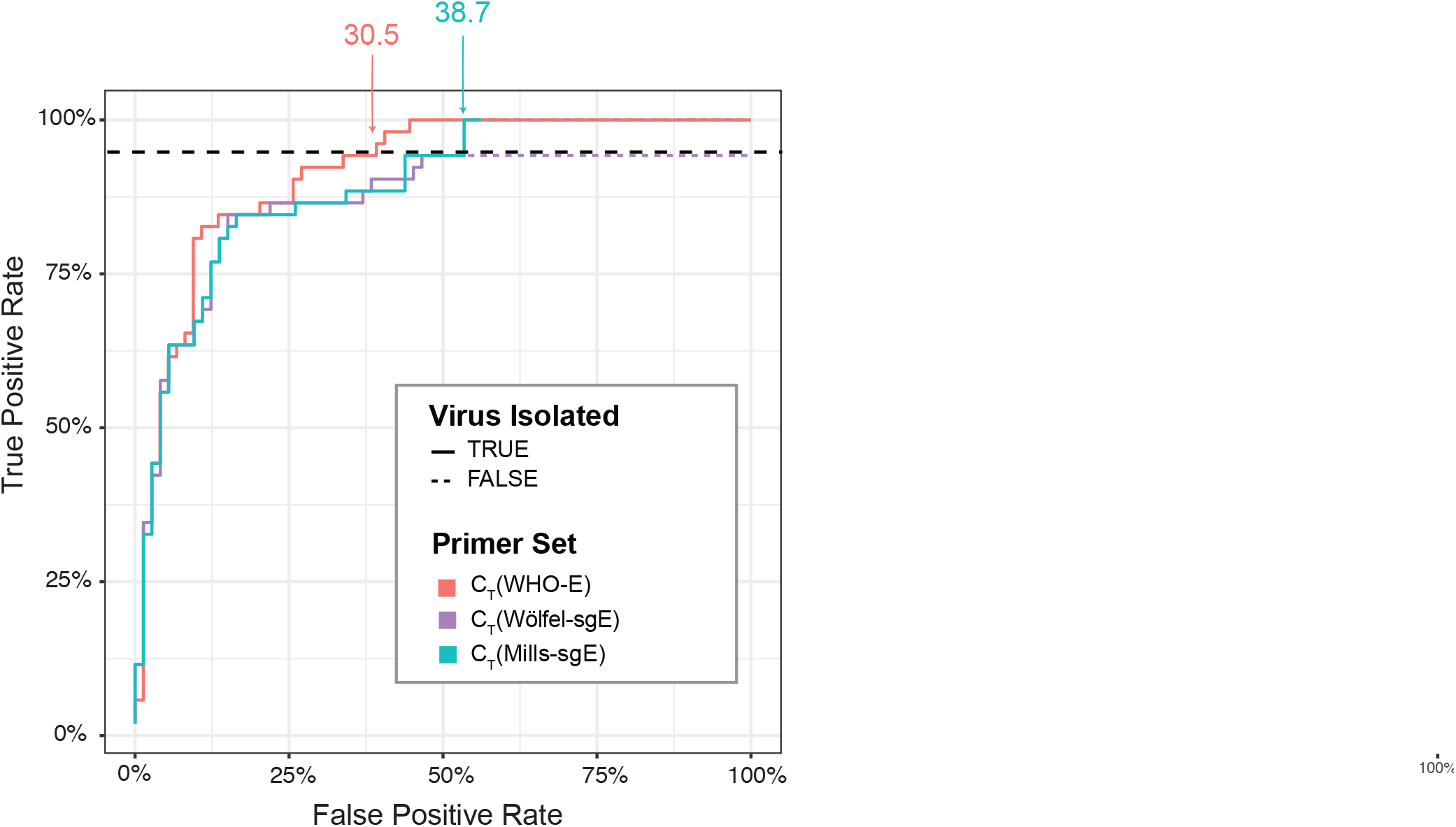
Receiver operating characteristic (ROC) curve comparing diagnostic performance of WHO-E, Wölfel-sgE and Mills-sgE primer-probe sets. ROC curves showing the ability to determine infectivity (as assayed by viral culture in Vero E6-TMPRSS2 cells) using the C_T_ values of WHO-E, Wölfel-sgE, and Mills-sgE for each of the samples for which these values were determined. For each test, the true-positive rate (chance of correctly predicting samples to be culturable at a given or lower C_T_) was plotted against the false-positive rate (chance of incorrectly predicting samples to be culturable at a given or lower C_T_) as a step function line. The lines show performance of the tests as the (C_T_) threshold for a positive result is increased; the lines are solid for detectable values and dashed when a primer set returned an undetectable value. Each line is labelled with the lowest C_T_ value that surpassed a 95% sensitivity threshold (the horizontal dashed line) for that marker.

### Presence of sgE in purified virions

While we were initially surprised by the number of samples with viral RNA detectable by Mills-sgE and Wölfel-sgE that lacked isolatable virus, and by our observation of sgRNA outside of cells, there are several potential explanations for this. First, RNA could be released into the supernatant by infected cells undergoing apoptosis. Second, given that sgE:E in the supernatants remain relatively constant (including at timepoints before cell death), it is also possible that SARS-CoV-2 sgRNA is packaged into virions and actively released. While not yet identified for SARS-CoV-2, it is generally thought that packaging signals within the coronavirus genome control the specific incorporation of genomic RNA and exclude sgRNA species [28-31]; other reports suggest that subgenomic RNAs can be packaged and may serve as a template for early replication [32, 33]. To investigate this possibility, we obtained virus concentrated through a sucrose cushion by ultracentrifugation and compared this to RNA from infected cells or microcentrifuge-clarified supernatants from those same cells (Fig. 8). Relative to the total amount of E RNA within each sample type, both a cellular control gene (GAPDH) and negative-strand E were found at the highest levels in cells, with decreasing abundance in clarified supernatants and ultracentrifuge concentrated samples respectively. The low levels of GAPDH seen in the samples concentrated by ultracentrifuge (less than 0.1% of levels seen in cellular lysates) in particular suggests that these samples are enriched for released SARS-CoV-2 virions. Intriguingly, we observed very similar amounts of sgE (relative to E) in concentrated virus and clarified supernatant, suggesting the subgenomic RNA seen outside the cell may be specifically packaged in virions rather than simply associated with cellular debris.

**Figure 8.**
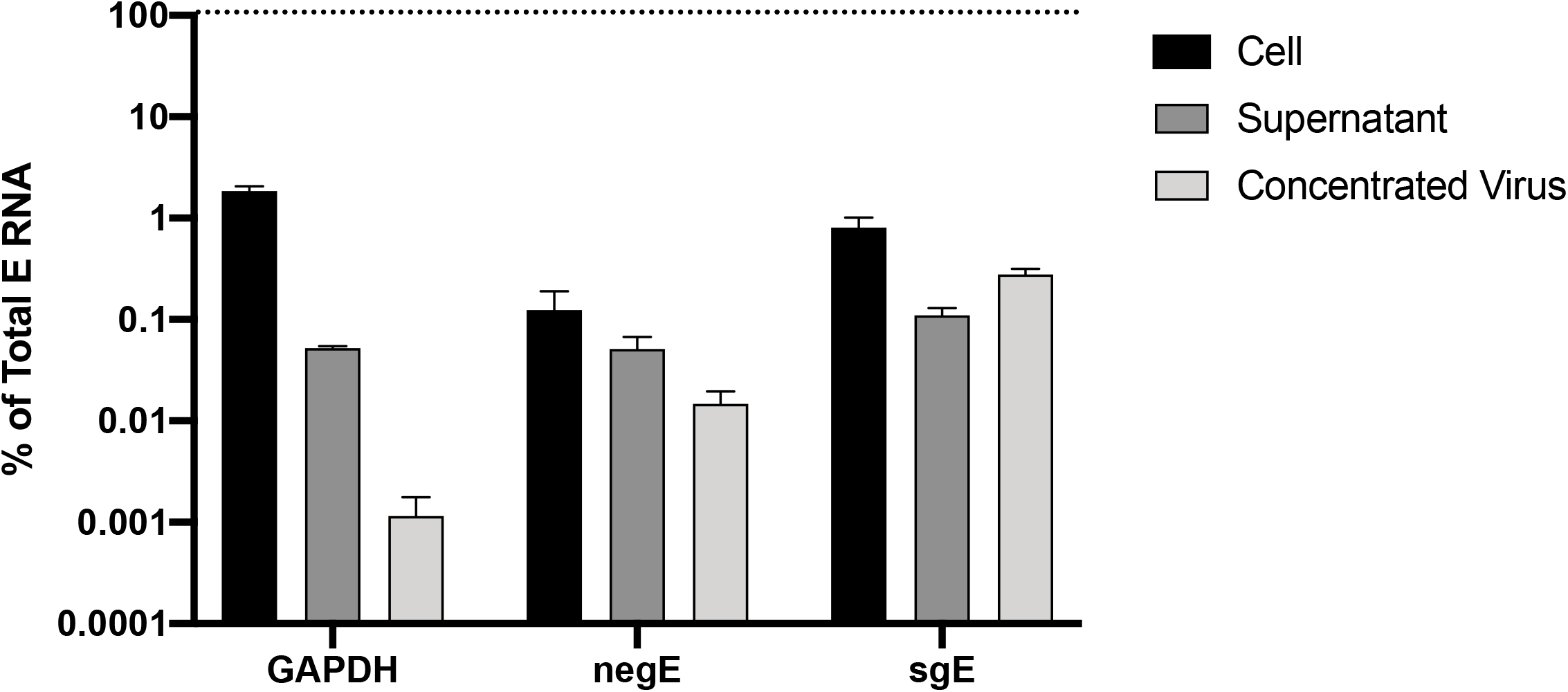
Relative abundance of sgE, negative-strand E, and GAPDH in cells, supernatants and purified virus. Levels of the cellular housekeeping gene GAPDH, negative-strand E (negE), and sgE relative to total levels of E RNA were measured in infected cells, clarified supernatants (4 dpi Vero-E6 samples from Fig. 2) and virus concentrated through a sucrose cushion by ultracentrifugation. Data points represent four technical replicates from two independent experiments (Cell and Supernatant) or three replicates (Concentrated Virus) and are normalized to the total levels of E for each sample, plotted as mean with error bars plotted as +/-SEM.

## DISCUSSION

Here, we show that using a threshold Ct (>31) using WHO-E or detection of sgRNA offered 100% negative predictive value for the likelihood of culturable virus being present in a given clinical specimen. Our data add to the growing body of knowledge that indicates quantitative viral load thresholds or subgenomic RNA testing serve as potential molecular correlates of infectivity beyond qualitative qRT-PCR tests that detect low levels of genomic RNA [34]. Our results and others [35] indicate that sgRNA is relatively stable and does not degrade more quickly than genomic RNA after active replication has ceased, and so while the absence of subgenomic RNA could be useful as a rule-out test, the presence of subgenomic RNA is not itself a marker for the presence of infectious virus or active infection.

While not unexpected, the findings that more viral RNA leads to an increased likelihood of viral isolation, a faster induction of CPE, and an infection that reaches peak titer more quickly, all underscore the importance of minimizing exposure wherever possible. Infection with SARS-CoV-2 is not a binary event, as more virus results in a faster infection while less virus results in delayed growth kinetics. At least in the setting of tissue culture, this may have important implications for the induction of the interferon response [36]. It is also important to note that the reporting threshold for the presence of infectious virus needs to take cell type into account. As most current isolation studies have been performed in Vero E6 cells [4, 5, 37] it is likely the viral RNA level at which infectious virus was presumed to be absent was artificially high, as in our hands many samples that did not isolate in Vero E6 cells successfully isolated in Vero E6-TMPRSS2 cells. While we observed that successful viral isolation was heavily dependent on starting RNA level, we were unable to isolate virus from several samples with very low C_T_ levels, likely reflecting the unknown storage conditions (precise temperatures, time from sample collection to processing, etc.) typical of clinical samples, and particularly those collected during a pandemic.

There are a number of limitations to the work presented here. Most importantly, it is not known whether viral isolation is a perfect laboratory correlate for viral infectivity and transmission in humans, which can vary significantly by time, distance, anatomy or mask wearing, and host immune status. We did not specifically convert CT to viral load given the multiple loci and tests investigated and the presence of mixed genomic and subgenomic transcripts for certain qRT-PCR sets. C_T_ values are strongly assay-and instrument-dependent, and so other labs would need to validate the sensitivity of these primers against independent standard curves in order to calibrate assay performance before putting either the WHO-E C_T_ limit or the Mills-sgE assay into use in their own labs.

Finally, this work raises several important questions regarding the basic virology of SARS-CoV-2. The variation in viral titers generated from samples harvested at similar levels of CPE is intriguing, especially as we observed relatively little variation in RNA levels seen in these same samples (Supplemental Table S2). Potential variations in the ratio of infectious virus titers to RNA levels has important implications, as current dogma generally assumes a constant relationship between total RNA and levels of infectious virus in clinical samples. Higher levels of viral RNA correlate with poorer clinical outcomes for instance [38], but in general it has been difficult if not impossible to routinely measure infectious titers directly in clinical samples.

The presence of abundant sgRNA in viral supernatants at timepoints before cell lysis and also in concentrated virions was also surprising, suggesting that these species may be packaged into virions, or alternatively, released into the supernatant through an alternative pathway. However, virus concentration through a sucrose cushion is not equivalent to highly purified virus (ie, sucrose banding), and will presumably include other extracellular vesicles such as exosomes. It remains an open question if sgRNA is still detected in highly purified virions, and whether this changes at any point during infection. The stability of viral RNA for many days after the near complete loss of infectivity may suggest an explanation for how RNA is detected for so long after initial infection in some patients. However, reports from immunocompromised patients show that active replication can undergo a cyclical progression [6] and the source of RNA detected weeks or months after infection remains to be fully elucidated. Given emerging prevalence of ‘long haulers’ with persistent symptoms, it is crucial to further investigate when and where viral replication takes place, and how common a pattern of cyclical replication is in patients. Finally, the subgenomic RNA assay described here is also suitable for use in specialized testing situations, such as distinguishing active infection from input viral inoculum in animal or vaccine trials [15] or from nucleic acid contamination of scientists working with SARS-CoV-2 plasmids leading to false positive screening tests [39, 40].

## METHODS

### Viruses and cells

The use of deidentified positive specimens for the above studies was approved by the University of Washington Institutional Review Board (STUDY00010205). SARS-CoV-2 viral isolations and growth curves were conducted at the UVM BSL-3 facility under an approved Institutional Biosafety protocol. SARS-CoV-2 strain 2019-nCoV/USA_USA-WA1/2020 (WA1) was generously provided by Kenneth Plante and the World Reference Center for Emerging Viruses and Arboviruses at the University of Texas Medical Branch and propagated in African green monkey kidney cells (Vero E6) that were kindly provided by J.L Whitton. Viral RNA (courtesy of David Bauer, The Francis Crick Institute, UK) from concentrated SARS-CoV-2 (England02 strain, B lineage ‘Wuhan-like’) was obtained by clarifying viral supernatants, pelleting overnight in a floor standing centrifuge, resuspending and purifying by ultracentrifugation through a sucrose cushion. Pellets were then resuspended in buffer and extracted with TRIzol LS. Vero E6 cells expressing the TMPRSS-2 protease were obtained from the JCRB Cell Bank (JCRB No. JCRB1819). Vero E6 cells were maintained in complete Dulbecco’s Modified Eagle Medium (cDMEM; Thermo Fisher, Cat. #11965–092) containing 10% fetal bovine serum (Gibco, Thermo-Fisher, Cat. #16140–071), 1% HEPES Buffer Solution (15630–130), and 1% penicillin– streptomycin (Thermo Fisher, Cat. #15140–122). Vero E6-TMPRSS2 cells were maintained in the same media with the addition of G418. Cells were grown in a humidified incubator at 37°C with 5% CO_2_.

### Viral isolations

Nasopharyngeal patient samples previously verified as SARS-CoV-2 positive by the University of Washington Virology Laboratory were used for viral isolations and viral RNA measurements. Each sample was frozen once at -80°C before viral growth experiments. Vero E6 or Vero E6-TMPRSS2 cells were seeded in diagonally adjacent wells of 24 well plates (12 wells seeded/plate to minimize cross contamination risk) at 3.5⨯10^5^ cells/well one day prior to infecting. 100 µL of each clinical sample was used to inoculate parallel Vero E6 or Vero E6-TMPRSS2 monolayers, for 1 hour at 37°C with rocking. After the 1 hr incubation, wells were individually aspirated, washed with PBS and overlaid with 1 ml standard Vero media containing 2% FBS. Wells were monitored daily and 100 µL of media was removed each day for subsequent RNA extraction. When unambiguous CPE was observed, cells were harvested and lysates clarified for subsequent RNA extraction and focus forming assays. Cells were suspended in RLT buffer (Qiagen) and viral supernatants were mixed 1:1 with AVE Viral Lysis Buffer (Qiagen) before RNA extraction.

### Focus forming assay

Viral titer was determined by focus forming assay in a 96 well plate format. Serial ten-fold dilutions of clarified viral supernatants were used to inoculate Vero E6 cell monolayers (60,000 cells/well seeded one day prior) in 96□well white polystyrene microplates (Thermo Fisher, Cat. #07-200-628). 50 µL of each virus dilution was inoculated onto the cells and incubated at 37°C in a 5% CO_2_ incubator for 60 min, after which the wells were overlaid with 1.2% methylcellulose in DMEM and incubated at 37°C in a 5% CO_2_ incubator for 24 h. Infected cells were fixed in 25% formaldehyde in 3 × PBS. Cells were permeabilized with 0.1% 100X Triton in 1× PBS for 15 min and then incubated with a primary, cross-reactive rabbit anti-SARS-CoV N monoclonal antibody (Sinobiological, distributed by Thermo Fisher, Cat. #40143-R001 at a dilution of 1:20,000) followed by a peroxidase-labelled goat anti-rabbit antibody (SeraCare, Milford, MA, USA, Cat. #5220-0336 diluted to 1:2,000) and then the peroxidase substrate (SeraCare, Cat. #5510-0030).

### RNA extractions

Total nucleic acid (TNA) in all clinical NP samples, the highest-concentration sample from the AccuPlex SARS-CoV-2 Verification Panel (Member 1: 100,000 copies/mL of synthetic whole SARS-CoV-2 genome, SeraCare, Cat. #0505-0168), and cell and supernatant samples from viral culture were all extracted with an automated guanidinium lysis/magnetic silica bead absorption method using either Roche MagNA Pure LC instrument and Total Nucleic Acid Isolation Kit – High Performance (Roche, Cat. #05 323 738 001) or MagNA Pure 96 instrument and DNA & Viral NA Small Volume kit (Roche, Cat. # 05 467 497 001) according to manufacturer instructions. All extractions used 200 µl of input volume and were eluted into 50 µl.

### RT-PCR

Specific viral RNA was reverse transcribed into cDNA and then amplified in real-time PCR reactions using AgPath-ID One Step RT-PCR kit (Life Technologies, ThermoFisher, Cat. #4387424M), using 5 µl of extracted TNA per 25 µl reaction. RT-PCR reactions used one of four sets of primers/probes: 1) WHO-E, using the E_Sarbecco-F/R/P set; 2) Wölfel-sgE, using sgLeader-F with E_Sarbecco-R/P; 3) Mills-sgE, using sgLeader-F2, sgE-R, and sgE-P; and 4) GAPDH, using the primer/probe set for Rhesus macaque (ThermoFisher, Cat. #4331182, Rh02621745_g1). RT-PCR was performed on an ABI 7500 real-time PCR system as previously described [41] (Table 3).

**Table 3.**
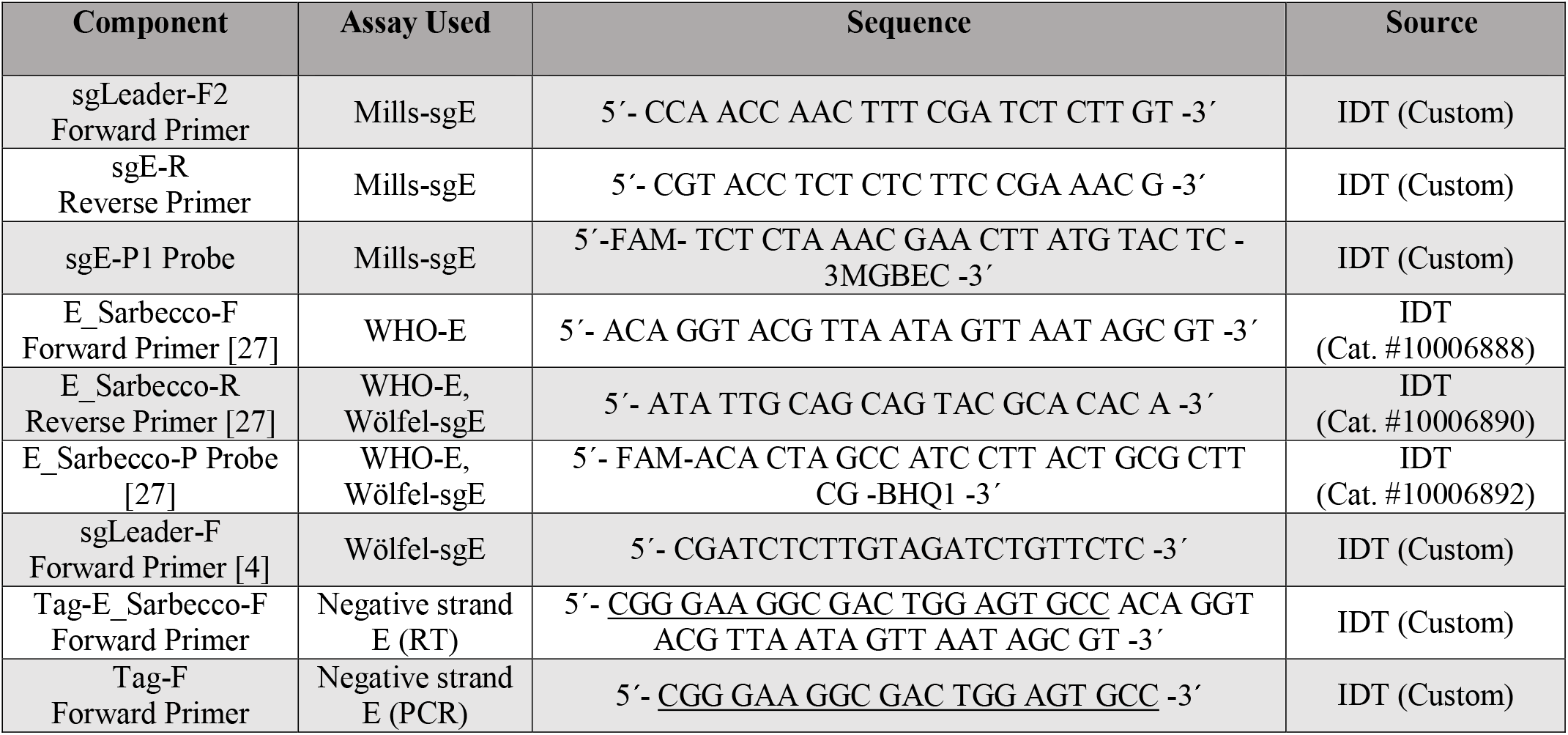
Primer/Probe Sequences.

### Negative-strand amplification

To avoid the nonspecific priming exhibited in reverse transcription of viral RNA, strand-specific amplification was accomplished in two steps, following the method of Vashist and colleagues [42]. First, viral negative-strand RNA was reverse transcribed with SuperScriptIII RT (Life Technologies, Cat. #18080-044) in half-reactions following manufacturer instructions, with 5 µl of extracted TNA in each 10 µl RT reaction. A primer containing a non-viral Tag sequence, Tag-E_Sarbecco-F, was used to prime the cDNA synthesis. Second, only cDNA resulting from the tagged RT reaction was amplified in real-time PCR reactions using QuantiTect Multiplex PCR kits. Each 30 µl PCR reaction contained 5 µl of cDNA, 4.7 µl water, 14.3 µl NoROX (Qiagen, Cat. #204745), 0.7 µl Hi Rox (Qiagen, Cat. #204545), 2.5 µl each of 10 uM Tag-F and E_Sarbecco-R primers, and 0.3 µl of 10 µM E_Sarbecco-P probe. PCR was performed on an ABI 7500 real-time PCR system with the following cycling parameters: 15’ at 95°C followed by 40 cycles of 60” at 94°C and 60” at 60°C (Table 3).

### sgE Transcript

A synthetic gBlock containing a T7 promoter, the SARS-CoV-2 genomic leader, TRS, and the first 154 bases of E gene was ordered from IDT. This gBlock was used as template for *in vitro* transcription reactions with HiScribe T7 RNA Synthesis Kit (New England Biolabs, Cat. #E2040S) following manufacturer instructions. RNA was transcribed for 16 hours at 37°C, then DNase treated and purified using illustra G-25 spin columns (GE Healthcare, Buckinghamshire, United Kingdom, Cat. #27532501). Concentration of the resulting RNA was determined first by NanoDrop spectrophotometer of two high-concentration dilutions (approximately 1 µg/µl and 100 ng/µl) measured in duplicate followed by a dilution in PBS to an approximate concentration of 2×10^11^ copies/mL, and then by reverse transcription droplet digital PCR (RT-ddPCR) system (Bio-Rad, Hercules, CA, USA) of two low-concentration dilutions (approximately 100 and 10 copies/µl) measured in duplicate with the Mills-sgE primer/probe set.

### ROC analysis

ROC curves were constructed and plotted using R [43].

## Data Availability

The data supporting the findings of this study are available within the manuscript and its supplementary materials. R code is available upon reasonable request.

## FIGURE LEGENDS

<u>Supplementary Figure 1. Receiver operating characteristic (ROC) curve comparing diagnostic performance of WHO-E, Wölfel-sgE, Mills-sgE and negative-strand E primer-probe sets.</u>

ROC curves showing the ability to determine infectivity (as assayed by viral culture in Vero E6-TMPRSS2 cells) using the C_T_ values of WHO-E, Wölfel-sgE, Mills-sgE, and negative-strand E for a subset of the samples from Fig 7 for which we had sufficient material to test these four primer sets. For each test, the true-positive rate (chance of correctly predicting samples to be culturable at a given or lower C_T_) was plotted against the false-positive rate (chance of incorrectly predicting samples to be culturable at a given or lower C_T_) as a step function line. The lines show performance of the tests as the (C_T_) threshold for a positive result is increased; the lines are solid for detectable values and dashed when a primer set returned an undetectable value. Each line is labelled with the lowest C_T_ value that surpassed a 95% sensitivity threshold (the horizontal dashed line) for that marker.

## ACKNOWLEDGMENTS

We are grateful to David Bauer and Saira Hussain for the kind gift of RNA from sucrose concentrated SARS-CoV-2. We thank Madaline Schmidt, Annalis Whitaker, Philip Eisenhauer, and Joyce Oetjen for technical assistance. This work was supported by NIH P30GM118228-04 (to E.A.B.) and funding from the Office of the Vice President for Research at the University of Vermont (JWB).

## CONFLICTS OF INTEREST

ALG reports contract testing from Abbot and research support from Merck and Gilead. The other authors declare no conflicts of interest.

## Notes

### Author Declarations

The excess clinical samples used in this study were approved under a waiver of consent by the University of Washington institutional review board (IRB; STUDY00000408). The de-identified samples were determined to be exempt because they were not considered human subjects research due to the quality improvement and public health intent of the work.

## REFERENCES

1. Walsh KA, Jordan K, Clyne B, Rohde D, Drummond L, Byrne P, et al. SARS-CoV-2 detection, viral load and infectivity over the course of an infection. J Infect. 2020;81(3):357–71. Epub 2020/07/03. doi: 10.1016/j.jinf.2020.06.067. PubMed PMID: 32615199; PubMed Central PMCID: PMCPMC7323671.

2. He X, Lau EHY, Wu P, Deng X, Wang J, Hao X, et al. Temporal dynamics in viral shedding and transmissibility of COVID-19. Nat Med. 2020;26(5):672–5. Epub 2020/04/17. doi: 10.1038/s41591-020-0869-5. PubMed PMID: 32296168.

3. Cevik M, Tate M, Lloyd O, Maraolo AE, Schafers J, Ho A. SARS-CoV-2, SARS-CoV, and MERS-CoV viral load dynamics, duration of viral shedding, and infectiousness: a systematic review and meta-analysis. The Lancet Microbe. doi: 10.1016/S2666-5247(20)30172-5.

4. Wolfel R, Corman VM, Guggemos W, Seilmaier M, Zange S, Muller MA, et al. Virological assessment of hospitalized patients with COVID-2019. Nature. 2020. Epub 2020/04/03. doi: 10.1038/s41586-020-2196-x. PubMed PMID: 32235945.

5. van Kampen JJA, van de Vijver DAMC, Fraaij PLA, Haagmans BL, Lamers MM, Okba N, et al. Shedding of infectious virus in hospitalized patients with coronavirus disease-2019 (COVID-19): duration and key determinants. medRxiv. 2020:2020.06.08.20125310. doi: 10.1101/2020.06.08.20125310.

6. Baang JH, Smith C, Mirabelli C, Valesano AL, Manthei DM, Bachman M, et al. Prolonged SARS-CoV-2 replication in an immunocompromised patient. J Infect Dis. 2020. Epub 2020/10/23. doi: 10.1093/infdis/jiaa666. PubMed PMID: 33089317.

7. Lai MM, Cavanagh D. The molecular biology of coronaviruses. Adv Virus Res. 1997;48:1–100. Epub 1997/01/01. doi: 10.1016/s0065-3527(08)60286-9. PubMed PMID: 9233431; PubMed Central PMCID: PMCPMC7130985.

8. Sawicki SG, Sawicki DL. A new model for coronavirus transcription. Adv Exp Med Biol. 1998;440:215–9. Epub 1998/10/23. doi: 10.1007/978-1-4615-5331-1_26. PubMed PMID: 9782283.

9. Speranza E, Williamson BN, Feldmann F, Sturdevant GL, Pérez-Pérez L, Mead-White K, et al. SARS-CoV-2 infection dynamics in lungs of African green monkeys. bioRxiv. 2020. Epub 2020/08/26. doi: 10.1101/2020.08.20.258087. PubMed PMID: 32839775; PubMed Central PMCID: PMCPMC7444286.

10. Wong CH, Ngan CY, Goldfeder RL, Idol J, Kuhlberg C, Maurya R, et al. Subgenomic RNAs as molecular indicators of asymptomatic SARS-CoV-2 infection. bioRxiv. 2021:2021.02.06.430041. doi: 10.1101/2021.02.06.430041.

11. Zollo M, Ferrucci V, Izzo B, Quarantelli F, Domenico CD, Comegna M, et al. SARS-CoV-2 Subgenomic N (sgN) Transcripts in Oro-Nasopharyngeal Swabs Correlate with the Highest Viral Load, as Evaluated by Five Different Molecular Methods. Diagnostics (Basel). 2021;11(2). Epub 2021/03/07. doi: 10.3390/diagnostics11020288. PubMed PMID: 33673182; PubMed Central PMCID: PMCPMC7923082.

12. Haberman M, Wu, K.J., Mandavilli, A. White House Doctor Says Trump Is No Longer Contagious. The New York Times. 2020 10/10/2020.

13. Perera R, Tso E, Tsang OTY, Tsang DNC, Fung K, Leung YWY, et al. SARS-CoV-2 Virus Culture and Subgenomic RNA for Respiratory Specimens from Patients with Mild Coronavirus Disease. Emerg Infect Dis. 2020;26(11):2701–4. Epub 2020/08/05. doi: 10.3201/eid2611.203219. PubMed PMID: 32749957; PubMed Central PMCID: PMCPMC7588524.

14. Verma R, Kim E, Martinez-Colón GJ, Jagannathan P, Rustagi A, Parsonnet J, et al. SARS-CoV-2 subgenomic RNA kinetics in longitudinal clinical samples. Open Forum Infectious Diseases. 2021. doi: 10.1093/ofid/ofab310.

15. Dagotto G, Mercado NB, Martinez DR, Hou YJ, Nkolola JP, Carnahan RH, et al. Comparison of Subgenomic and Total RNA in SARS-CoV-2 Challenged Rhesus Macaques. Journal of Virology. 2021:pJVI.02370-20. doi: 10.1128/jvi.02370-20.

16. Mercado NB, Zahn R, Wegmann F, Loos C, Chandrashekar A, Yu J, et al. Single-shot Ad26 vaccine protects against SARS-CoV-2 in rhesus macaques. Nature. 2020;586(7830):583–8. Epub 2020/07/31. doi: 10.1038/s41586-020-2607-z. PubMed PMID: 32731257; PubMed Central PMCID: PMCPMC7581548.

17. Chandrashekar A, Liu J, Martinot AJ, McMahan K, Mercado NB, Peter L, et al. SARS-CoV-2 infection protects against rechallenge in rhesus macaques. Science. 2020;369(6505):812–7. Epub 2020/05/22. doi: 10.1126/science.abc4776. PubMed PMID: 32434946; PubMed Central PMCID: PMCPMC7243369.

18. van Doremalen N, Lambe T, Spencer A, Belij-Rammerstorfer S, Purushotham JN, Port JR, et al. ChAdOx1 nCoV-19 vaccine prevents SARS-CoV-2 pneumonia in rhesus macaques. Nature. 2020;586(7830):578–82. Epub 2020/07/31. doi: 10.1038/s41586-020-2608-y. PubMed PMID: 32731258.

19. Corbett KS, Flynn B, Foulds KE, Francica JR, Boyoglu-Barnum S, Werner AP, et al. Evaluation of the mRNA-1273 Vaccine against SARS-CoV-2 in Nonhuman Primates. New England Journal of Medicine. 2020;383(16):1544–55. doi: 10.1056/NEJMoa2024671.

20. Hogan CA, Huang C, Sahoo MK, Wang H, Jiang B, Sibai M, et al. Strand-Specific Reverse Transcription PCR for Detection of Replicating SARS-CoV-2. Emerging Infectious Diseases. 2021;27:632–5. doi: https://dx.doi.org/10.3201/eid2702.204168.

21. Hoffmann M, Kleine-Weber H, Schroeder S, Krüger N, Herrler T, Erichsen S, et al. SARS-CoV-2 Cell Entry Depends on ACE2 and TMPRSS2 and Is Blocked by a Clinically Proven Protease Inhibitor. Cell. 2020;181(2):271-80.e8. Epub 2020/03/07. doi: 10.1016/j.cell.2020.02.052. PubMed PMID: 32142651; PubMed Central PMCID: PMCPMC7102627.

22. Hoffmann M, Mösbauer K, Hofmann-Winkler H, Kaul A, Kleine-Weber H, Krüger N, et al. Chloroquine does not inhibit infection of human lung cells with SARS-CoV-2. Nature. 2020;585(7826):588–90. Epub 2020/07/23. doi: 10.1038/s41586-020-2575-3. PubMed PMID: 32698190.

23. Bullard J, Dust K, Funk D, Strong JE, Alexander D, Garnett L, et al. Predicting infectious SARS-CoV-2 from diagnostic samples. Clin Infect Dis. 2020. Epub 2020/05/23. doi: 10.1093/cid/ciaa638. PubMed PMID: 32442256; PubMed Central PMCID: PMCPMC7314198.

24. Gniazdowski V, Morris CP, Wohl S, Mehoke T, Ramakrishnan S, Thielen P, et al. Repeat COVID-19 Molecular Testing: Correlation of SARS-CoV-2 Culture with Molecular Assays and Cycle Thresholds. Clin Infect Dis. 2020. Epub 2020/10/27. doi: 10.1093/cid/ciaa1616. PubMed PMID: 33104776; PubMed Central PMCID: PMCPMC7665437.

25. Jaafar R, Aherfi S, Wurtz N, Grimaldier C, Hoang VT, Colson P, et al. Correlation between 3790 qPCR positives samples and positive cell cultures including 1941 SARS-CoV-2 isolates. Clin Infect Dis. 2020. Epub 2020/09/29. doi: 10.1093/cid/ciaa1491. PubMed PMID: 32986798; PubMed Central PMCID: PMCPMC7543373.

26. Singanayagam A, Patel M, Charlett A, Lopez Bernal J, Saliba V, Ellis J, et al. Duration of infectiousness and correlation with RT-PCR cycle threshold values in cases of COVID-19, England, January to May 2020. Euro Surveill. 2020;25(32). Epub 2020/08/15. doi: 10.2807/1560-7917.Es.2020.25.32.2001483. PubMed PMID: 32794447; PubMed Central PMCID: PMCPMC7427302.

27. Corman VM, Landt O, Kaiser M, Molenkamp R, Meijer A, Chu DKW, et al. Detection of 2019 novel coronavirus (2019-nCoV) by real-time RT-PCR. Euro Surveill. 2020;25 (3). Epub 2020/01/30. doi: 10.2807/1560-7917.ES.2020.25.3.2000045. PubMed PMID: 31992387; PubMed Central PMCID: PMCPMC6988269.

28. Makino S, Yokomori K, Lai MM. Analysis of efficiently packaged defective interfering RNAs of murine coronavirus: localization of a possible RNA-packaging signal. J Virol. 1990;64(12):6045–53. Epub 1990/12/01. doi: 10.1128/jvi.64.12.6045-6053.1990. PubMed PMID: 2243386; PubMed Central PMCID: PMCPMC248778.

29. Escors D, Izeta A, Capiscol C, Enjuanes L. Transmissible gastroenteritis coronavirus packaging signal is located at the 5’ end of the virus genome. J Virol. 2003;77(14):7890–902. Epub 2003/06/28. doi: 10.1128/jvi.77.14.7890-7902.2003. PubMed PMID: 12829829; PubMed Central PMCID: PMCPMC161917.

30. Kuo L, Masters PS. Functional analysis of the murine coronavirus genomic RNA packaging signal. J Virol. 2013;87(9):5182–92. Epub 2013/03/02. doi: 10.1128/jvi.00100-13. PubMed PMID: 23449786; PubMed Central PMCID: PMCPMC3624306.

31. Athmer J, Fehr AR, Grunewald ME, Qu W, Wheeler DL, Graepel KW, et al. Selective Packaging in Murine Coronavirus Promotes Virulence by Limiting Type I Interferon Responses. mBio. 2018;9(3):e00272–18. doi: 10.1128/mBio.00272-18.

32. Hofmann MA, Sethna PB, Brian DA. Bovine coronavirus mRNA replication continues throughout persistent infection in cell culture. J Virol. 1990;64(9):4108–14. Epub 1990/09/01. doi: 10.1128/jvi.64.9.4108-4114.1990. PubMed PMID: 2384915; PubMed Central PMCID: PMCPMC247873.

33. Zhao X, Shaw K, Cavanagh D. Presence of subgenomic mRNAs in virions of coronavirus IBV. Virology. 1993;196(1):172–8. Epub 1993/09/01. doi: 10.1006/viro.1993.1465. PubMed PMID: 8395112; PubMed Central PMCID: PMCPMC7130481.

34. Jefferson T, Spencer EA, Brassey J, Heneghan C. Viral cultures for COVID-19 infectious potential assessment - a systematic review. Clin Infect Dis. 2020. Epub 2020/12/04. doi: 10.1093/cid/ciaa1764. PubMed PMID: 33270107; PubMed Central PMCID: PMCPMC7799320.

35. Alexandersen S, Chamings A, Bhatta TR. SARS-CoV-2 genomic and subgenomic RNAs in diagnostic samples are not an indicator of active replication. Nature Communications. 2020;11(1):6059. doi: 10.1038/s41467-020-19883-7.

36. Blanco-Melo D, Nilsson-Payant BE, Liu WC, Uhl S, Hoagland D, Møller R, et al. Imbalanced Host Response to SARS-CoV-2 Drives Development of COVID-19. Cell. 2020;181(5):1036-45.e9. Epub 2020/05/18. doi: 10.1016/j.cell.2020.04.026. PubMed PMID: 32416070; PubMed Central PMCID: PMCPMC7227586.

37. Arons MM, Hatfield KM, Reddy SC, Kimball A, James A, Jacobs JR, et al. Presymptomatic SARS-CoV-2 Infections and Transmission in a Skilled Nursing Facility. N Engl J Med. 2020. Epub 2020/04/25. doi: 10.1056/NEJMoa2008457. PubMed PMID: 32329971.

38. Bryan A, Fink SL, Gattuso MA, Pepper G, Chaudhary A, Wener MH, et al. SARS-CoV-2 Viral Load on Admission Is Associated With 30-Day Mortality. Open Forum Infectious Diseases. 2020;7(12). doi: 10.1093/ofid/ofaa535.

39. Robinson-McCarthy LR, Mijalis AJ, Filsinger GT, de Puig H, Donghia NM, Schaus TE, et al. Anomalous COVID-19 tests hinder researchers. Science. 2021;371(6526):244–5. doi: 10.1126/science.abf8873.

40. Montgomery TL, Paavola M, Bruce EA, Botten JW, Crothers JW, Krementsov DN. Laboratory worker self-contamination with non-infectious SARS-CoV-2 DNA can result in false positive RT-PCR-based surveillance testing. J Clin Microbiol. 2021. Epub 2021/05/07. doi: 10.1128/JCM.00723-21. PubMed PMID: 33952597.

41. Nalla AK, Casto AM, Huang MW, Perchetti GA, Sampoleo R, Shrestha L, et al. Comparative Performance of SARS-CoV-2 Detection Assays using Seven Different Primer/Probe Sets and One Assay Kit. J Clin Microbiol. 2020. Epub 2020/04/10. doi: 10.1128/JCM.00557-20. PubMed PMID: 32269100.

42. Vashist S, Urena L, Goodfellow I. Development of a strand specific real-time RT-qPCR assay for the detection and quantitation of murine norovirus RNA. Journal of Virological Methods. 2012;184(1):69–76. doi: https://doi.org/10.1016/j.jviromet.2012.05.012.

43. R Development Core Team. R: A language and environment for statistical computing. Vienna, Austria: R Foundation for Statistical Computing; 2018.

