## Supplementary figures and images for "Predicting Infectivity: Comparing Four PCR-based Assays to Detect Culturable SARS-CoV-2 in Clinical Samples"

### Supplemental Figure 1

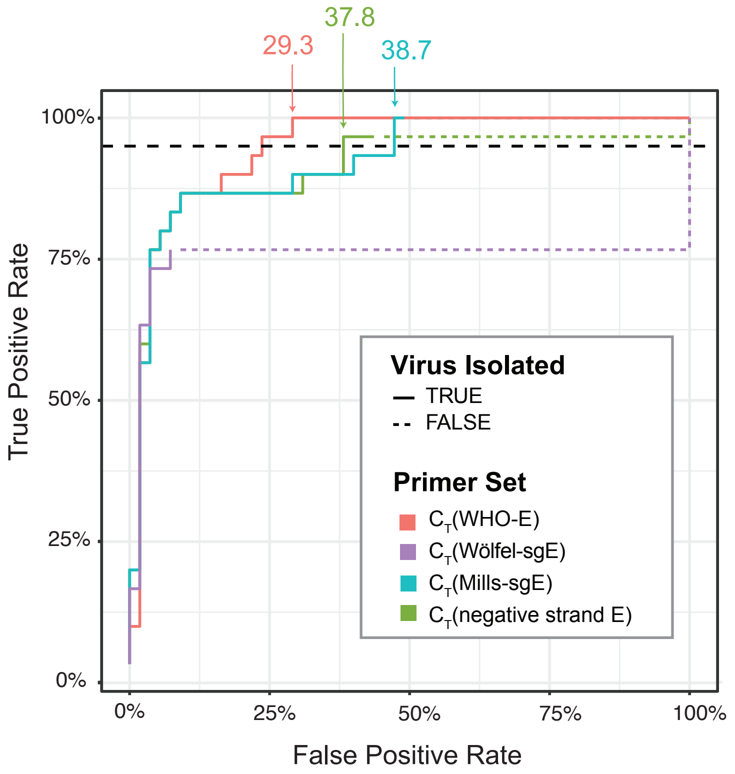
